# Genetic Diversity and Expanded Phenotypes in Dystonia: Insights from Large-Scale Exome Sequencing

**DOI:** 10.1101/2024.12.02.24316741

**Authors:** Mirja Thomsen, Fabian Ott, Sebastian Loens, Gamze Kilic-Berkmen, Ai Huey Tan, Shen-Yang Lim, Ebba Lohmann, Kaja M. Schröder, Lea Ipsen, Lena A. Nothacker, Linn Welzel, Alexandra S. Rudnik, Frauke Hinrichs, Thorsten Odorfer, Kirsten E. Zeuner, Friederike Schumann, Andrea A. Kühn, Simone Zittel, Marius Moeller, Robert Pfister, Christoph Kamm, Anthony E. Lang, Yi Wen Tay, Marie Vidailhet, Emmanuel Roze, Joel S. Perlmutter, Jeanne S. Feuerstein, Victor S. C. Fung, Florence Chang, Richard L. Barbano, Steven Bellows, Aparna A. Wagle Shukla, Alberto J. Espay, Mark S. LeDoux, Brian D. Berman, Stephen Reich, Andres Deik, Andre Franke, Michael Wittig, Sören Franzenburg, Jens Volkmann, Norbert Brüggemann, H. A. Jinnah, Tobias Bäumer, Christine Klein, Hauke Busch, Katja Lohmann

## Abstract

Dystonia is one of the most prevalent movement disorders, characterized by significant clinical and etiological heterogeneity. Despite considerable heritability (∼25%) and the identification of several disease-linked genes, the etiology in most patients remains elusive. Moreover, understanding the correlations between clinical manifestation and genetic variants has become increasingly complex.

To comprehensively unravel dystonia’s genetic spectrum, we performed exome sequencing on 1,924 dystonia patients [40.3% male, 92.9% White, 93.2% isolated dystonia, median age at onset (AAO) 33 years], including 1,895 index patients, who were previously genetically unsolved. The sample was mainly based on two dystonia registries (DysTract and the Dystonia Coalition). Further, 72 additional patients of Asian ethnicity, mainly from Malaysia, were also included. We prioritized patients with negative genetic prescreening, early AAO, positive family history, and multisite involvement of dystonia. Rare variants in genes previously linked to dystonia (*n*=405) were examined. Variants were confirmed via Sanger sequencing, and segregation analysis was performed when possible.

We identified 137 distinct likely pathogenic or pathogenic variants (according to ACMG criteria) across 51 genes in 163/1,924 patients [42.9% male, 85.9% White, 68.7% isolated dystonia, median AAO 19 years]. This included 153/1,895 index patients, resulting in a diagnostic yield of 8.1%. Notably, 77/137 (56.2%) of these variants were novel, with recurrent variants in *EIF2AK2*, *VPS16*, *KCNMA1*, and *SLC2A1*, and novel variant types such as two splice site variants in *KMT2B*, supported by functional evidence. Additionally, 321 index patients (16.9%) harbored variants of uncertain significance in 102 genes. The most frequently implicated genes included *VPS16*, *THAP1*, *GCH1*, *SGCE*, *GNAL*, and *KMT2B.* Presumably pathogenic variants in less well-established dystonia genes were also found, including *KCNMA1*, *KIF1A*, and *ZMYND11.* At least six variants (in *ADCY5*, *GNB1*, *IR2BPL, KCNN2*, *KMT2B*, and *VPS16*) occurred *de novo,* supporting pathogenicity. ROC curve analysis indicated that AAO and the presence of generalized dystonia were the strongest predictors of a genetic diagnosis, with diagnostic yields of 28.6% in patients with generalized dystonia and 20.4% in those with AAO < 30 years.

This study provides a comprehensive examination of the genetic landscape of dystonia, revealing valuable insights into the frequency of dystonia-linked genes and their associated phenotypes. It underscores the utility of exome sequencing in establishing diagnoses within this heterogeneous condition. Despite prescreening, presumably pathogenic variants were identified in almost 10% of patients. Our findings reaffirm several dystonia candidate genes and expand the phenotypic spectrum of some of these genes to include prominent, sometimes isolated dystonia.

## Introduction

Dystonia is a neurological disorder characterized by involuntary muscle contractions that lead to abnormal postures and repetitive movements. It ranks among the most prevalent movement disorders.^1^ Its clinical manifestations span a broad spectrum, from isolated cases to those combined with other movement disorders like parkinsonism, myoclonus, or chorea, to patients with additional neurological or systemic features.^1^

An essential aspect of establishing a dystonia diagnosis and appropriate treatment lies in genetic testing. However, despite the considerable heritability of dystonia, i.e., the presence of other affected family members with dystonia in about 25% of patients,^2^ and the identification of several causal genes, a significant proportion of patients remain without a genetic diagnosis, particularly those with later-onset isolated dystonia confined to one body region (focal dystonia).^3,4^ Genetic testing is complicated by the highly heterogeneous nature of the disease, as demonstrated by various large-scale sequencing studies,^4–6^ the ever-expanding list of dystonia-linked genes, and the reliance on clinical diagnosis due to the absence of an established biomarker. Presently, the Movement Disorder Society Task Force for the Nomenclature of Genetic Movement Disorders (MDS Nomenclature Task Force) recognizes 53 genes as established dystonia genes, including 10 genes for isolated dystonia, 10 for combined dystonia, and 33 for dystonia with other neurological or systemic features.^7^ Additionally, numerous genes have been linked to dystonia as a feature of other neurological disorders, further contributing to the complexity of the disorder. With ongoing next-generation sequencing efforts, the list of candidate genes for dystonia continues to grow, with many awaiting replication by independent studies.^4,7–9^ Altogether, this results in more than 400 potential dystonia-linked genes.^10^

The phenotypic presentations of dystonia are diverse, with considerable clinical overlap observed between the different genetic forms.^11,12^ Instances of classical phenotypes expanding over time or a single gene causing multiple distinct phenotypes, a phenomenon known as pleiotropy, further highlight this variability, as is the case for *ATP1A3*.^13^ In this regard, an increasing number of genes, previously associated with other neurological conditions including developmental delay, epileptic encephalopathy, and ataxia, are now recognized to play a significant role in patients with prominent, sometimes isolated dystonia (e.g., *CACNA1A*, *EIF2AK2*, *GNAO1*, *GNB1*, *TCF20*).^8,14^

As the phenotypic and mutational spectrum of dystonia expands, newly identified genetic variants require careful evaluation to establish a causal gene-disease link. Guidelines developed by the American College of Medical Genetics and Genomics,^15^ incorporating information from segregation analysis, in-silico predictions, and population databases, among others, are instrumental in this task. For some genes, specific functional assays have been developed to assess the impact of variants on protein function, such as aberrant CpG methylation serving as a functional readout for evaluating variants in the dystonia gene *KMT2B*.^16^ The most compelling evidence for novel dystonia genes, however, will always come from replicating the findings in independent patients and families.

Given the rarity and genetic complexity of dystonia, large cohorts are indispensable for comprehensively unraveling its genetic spectrum. To achieve this, we performed exome sequencing on 1,924 dystonia patients, the majority of whom were White, but also included individuals from diverse ethnic backgrounds such as Asian, African, Latino, American Indian, and Ashkenazi Jewish. In contrast to other dystonia cohorts typically enriched with early-onset, symptom-complex patients, this cohort is unique in that it predominantly consists of adult-onset isolated dystonia patients, which more accurately reflect the majority of patients seen in epidemiological studies. We describe detailed phenotypic and genetic findings in more than 400 genes previously linked to dystonia, including established dystonia genes, genes associated with other neurological disorders that may present dystonia as a phenotype, and recently proposed candidate genes for dystonia. This large-scale dataset provides valuable insights into the frequency and phenotypic manifestations of different genetic forms of dystonia and will aid future variant interpretation and clinical diagnostics.

## Materials and methods

### Study population

A total of 1,967 samples were included in the exome sequencing study, comprising 1,950 patients with dystonia and 17 unaffected family members (parents). One sample failed sequencing, and 25 samples were removed after relationship analysis (Supplementary Methods) revealed that those sample IDs referred to identical patients recruited twice from different sites. Altogether, this yielded a total number of 1,924 patients with dystonia without a previous genetic diagnosis who were included in the final cohort.

The vast majority of the samples were recruited in the framework of two large dystonia registries, DysTract (https://www.isms.uni-luebeck.de/en/research/dystract/) and the Dystonia Coalition (https://www.dystoniacoalition.org/). These samples had undergone genetic prescreening. Prescreening comprised: 1) Hot spot screening for known pathogenic variants in dystonia genes in about 80% of the samples using the Global Screening Array (GSA V01, Illumina; with custom content) and 2) Gene panel sequencing in about a third of the samples, comprising the genes *ANO3*, *GCH1*, *GNAL*, *KMT2B*, *PRKR*A, *SGCE*, *THAP1*, and *TOR1A* (results published elsewhere^17,18^). Patients with negative prescreening, early age at onset, positive family history, or multisite involvement were prioritized when selecting samples from the dystonia registries. Patient samples were collected in movement disorder clinics across Europe, North America, and Australia. Additionally, 72 samples from patients of Asian ethnicity (primarily from Malaysia) were included. The cohort consisted of diverse ethnic backgrounds, including White (*n*=1,787), Asian (*n*=72), African American (*n*=32), Latino (*n*=18), Ashkenazi Jewish (*n*=7), American Indian (*n*=5), and Mixed (*n*=3) individuals. Patients were phenotyped by movement disorder specialists and secondary causes, including acquired dystonia, were excluded. Written informed consent was obtained from all participants prior to the genetic testing, and the study was approved by the ethics committee at the University of Lübeck (04-180). The patient IDs used in this study cannot be linked to individual patients by anyone outside the research group.

### Sequencing and data processing

Genomic DNA was extracted from peripheral blood samples, and exome sequencing was performed at the Competence Centre for Genomic Analysis in Kiel, Germany, using Illumina NovaSeq. Sequencing reads were preprocessed and aligned to the Hg38 reference genome. Variant calling and annotation were conducted using DeepVariant, GLnexus, and VEP, with further details provided in the Supplementary Methods.

#### Variant filtering

A comprehensive list of dystonia-linked genes was generated by systematically querying the PubMed database (search term: dystoni* [TITLE/ABSTRACT] AND (gene* OR genetic* OR mutation* OR mutated OR varia*) AND "english"[Language]) until April 2024, resulting in 405 distinct genes that have been linked to dystonic symptoms in the literature (Supplementary Table 1). In this study, we utilized this phenotype-driven candidate gene list to identify genetic causes in our dystonia sample.

We filtered out variants with the following criteria: (a) low variant allele frequency (<20%), (b) low sequencing depth (<10 reads), (c) synonymous variants, (d) minor allele frequency >0.05% in the gnomAD population database, (e) variants classified as (likely) benign in ClinVar, and (f) single heterozygous variants in recessive disease genes. For the established dystonia genes that have been assigned a DYT prefix by the MDS Nomenclature Task Force,^7^ the remaining variants underwent detailed individual evaluation, and their pathogenicity was assessed using ACMG standards and guidelines.^15^ For genes not classified as established dystonia genes by the Task Force, additional filtering was applied to further narrow down the large number of rare variants: filtering out single heterozygous loss-of-function (LoF) variants in genes with good tolerance for such variants (gnomAD pLI-score = 0), missense variants classified as likely benign by the alpha missense score,^19^ and variants with a low CADD Score (<10),^20^ before individual evaluation by ACMG standards.

Only likely pathogenic or pathogenic variants according to ACMG criteria were considered disease-causing and underwent validation through Sanger sequencing. Candidate variants were tested for segregation whenever the DNA of family members was available.

The relevant literature for each gene was carefully reviewed to evaluate the phenotypic and genetic information associated with each gene. We categorized the phenotype-genotype relationships for all presumably pathogenic variants into four categories: consistent, partially consistent, quite inconsistent, and unclear/unknown. The unclear/unknown category was used when detailed clinical information was unavailable or for dystonia candidate genes without an established gene-phenotype relationship.

#### Statistical analysis

Statistical analysis and visualization were performed in R version 4.3.2. The analysis utilized base R functions for statistical tests and data manipulation, as well as the pROC package for generating and analysing ROC curves and the ggplot2 package for creating visualizations.

For calculating diagnostic yield, patients were grouped based on clinical criteria, including age at onset (AAO: <30 years, 30-50 years, and >50 years), dystonia subtypes (focal, segmental/multifocal, generalized), family history (positive or negative), and the presence of additional features versus isolated dystonia. These groupings, except for AAO, were also used for ROC analysis and statistical comparisons using Fisher’s exact test. AAO was treated as a continuous variable for both the Mann-Whitney U test comparing resolved and unresolved patients and in the ROC curve analysis. This approach was chosen to capture clinically relevant differences. Missing data were handled by excluding cases with NA values during analysis. All statistical tests were two-sided, and a p-value < 0.05 was considered statistically significant.

### Episignature analysis for *KMT2B* variants

For patients with rare variants in *KMT2B* and a sufficient amount of DNA available (*n*=45), the functional effect was assessed by analysing the diesease-specific methylation pattern (“episignature”) in peripheral blood, using the Illumina MethylationEPIC BeadChip. The mean of the normalized methylation levels (mean(z)) and the coefficient of variation (CV = SD/ |mean|) were used as quantifiers (Supplementary Methods).

## Results

### Patient characteristics and diagnostic yield

After quality control and excluding duplicated samples, the investigated cohort comprised a total of 1,924 dystonia patients without a previous genetic diagnosis, including 1,895 index cases and 29 affected family members. Patients had a median age at examination (AAE) of 54 years (interquartile range (IQR), 43-65), a median age at onset (AAO) of 33 years (IQR, 22.5- 43.5), and included 1,148 (59.7%) females and 776 (40.3%) males (Table 1). Of the patients, 1,045 (54.3%) had focal, 549 (28.5%) segmental or multifocal, and 236 (12.3%) generalized dystonia (Table 1). Among the focal dystonia patients, cervical dystonia was the most common subtype, observed in 550 (28.6%) patients. About 93% of all patients had isolated dystonia, while in 130 patients (6.8%), additional neurological (other than tremor) or systemic features were reported, such as bradykinesia (*n*=41), ataxia (*n*=35), myoclonus (*n*=35), or neurodevelopmental features (*n*=12). A positive family history was reported in 561 index patients (29.6%, 101 unknown).

**Table 1.** Patient characteristics in the investigated dystonia sample by diagnostic outcome.

|  | All patients | Patients with diagnostic variant | Patients with VUS | Yet-unsolved patients |
| --- | --- | --- | --- | --- |
| **Total patients** | 1,924 | 163 | 323 | 1,438 |
| Index patients | 1,895 | 153 (8.1%) | 321 (16.9%) | 1,421 (75.0%) |
| **Age at examination** | 54 years (43-65) | 46 years (32.5-59.5) | 55 years (45-65) | 55 years (44-66) |
| **Age at onset**<br>(median, IQR) | 33 years (22.5-43.5) | 19 years (5.25-32.75) | 34 years (24-44) | 34 years (24-44) |
| **Sex** |  |  |  |  |
| Males | 776 (40.3%) | 70 (42.9%) | 121 (37.5%) | 585 (40.7%) |
| Females | 1,148 (59.7%) | 93 (57.1%) | 202 (65.5%) | 853 (59.3%) |
| **Family history<sup>a</sup>** |  |  |  |  |
| Positive | 561 (29.6%) | 51 (33.3%) | 73 (22.7%) | 437 (30.9%) |
| Negative | 1,233 (65.1%) | 93 (60.8%) | 187 (58.3%) | 953 (66.9%) |
| NA | 101 (5.3%) | 9 (5.9%) | 61 (19.0%) | 31 (2.2%) |
| **Type of dystonia** |  |  |  |  |
| Generalized | 236 (12.3%) | 73 (44.8%) | 30 (9.3%) | 133 (9.2%) |
| Segmental/multifocal | 549 (28.5%) | 36 (22.1%) | 92 (28.5%) | 421 (29.3%) |
| Focal | 1,045 (54.3%) | 53 (32.5%) | 193 (59.8%) | 799 (55.6%) |
| NA | 94 (4.9%) | 1 (0.6%) | 8 (2.5%) | 85 (5.9%) |
| **Isolated dystonia** | 1,794 (93.2%) | 112 (68.7%) | 308 (95.4%) | 1,374 (95.5%) |
| **Additional neurological features reported (other than tremor)** | 130 (6.8%) | 51 (31.3%) | 15 (4.6%) | 64 (4.5%) |

IQR: interquartile range, NA: information not available, VUS: variant of uncertain significance
<sup>a</sup>calculated for index patients only

Exome sequencing revealed likely pathogenic or pathogenic variants in dystonia-linked genes in 163 patients, including 153 out of 1,895 index patients (8.1% diagnostic yield), with an additional 321 index patients (16.9%) having variants of uncertain significance (VUS) in dystonia-linked genes (Figure 1A, 2, and 3, Supplementary Table 2 and 3). Patients with likely pathogenic or pathogenic variants had a median AAO of 19 years (IQR, 5.25-32.75), and included 93 females and 70 males (Table 1); 73 patients had generalized (73/163, 44.8%), 36 multifocal or segmental (22.1%), and 53 focal dystonia (32.5%). For 51 patients (31.3%), additional features were reported, including myoclonus (*n*=20), neurodevelopmental features (*n*=12), bradykinesia (*n*=6), ataxia (*n*=5), and chorea (*n*=4). Positive family history was reported in 51/153 index patients (33.3%, 9 unknown). Regarding ethnicity, 140 patients were White (85.9%), 2 African American, 3 American Indian, 10 Asian, and for 8 patients, ethnicity was unknown.

**Figure 1.**
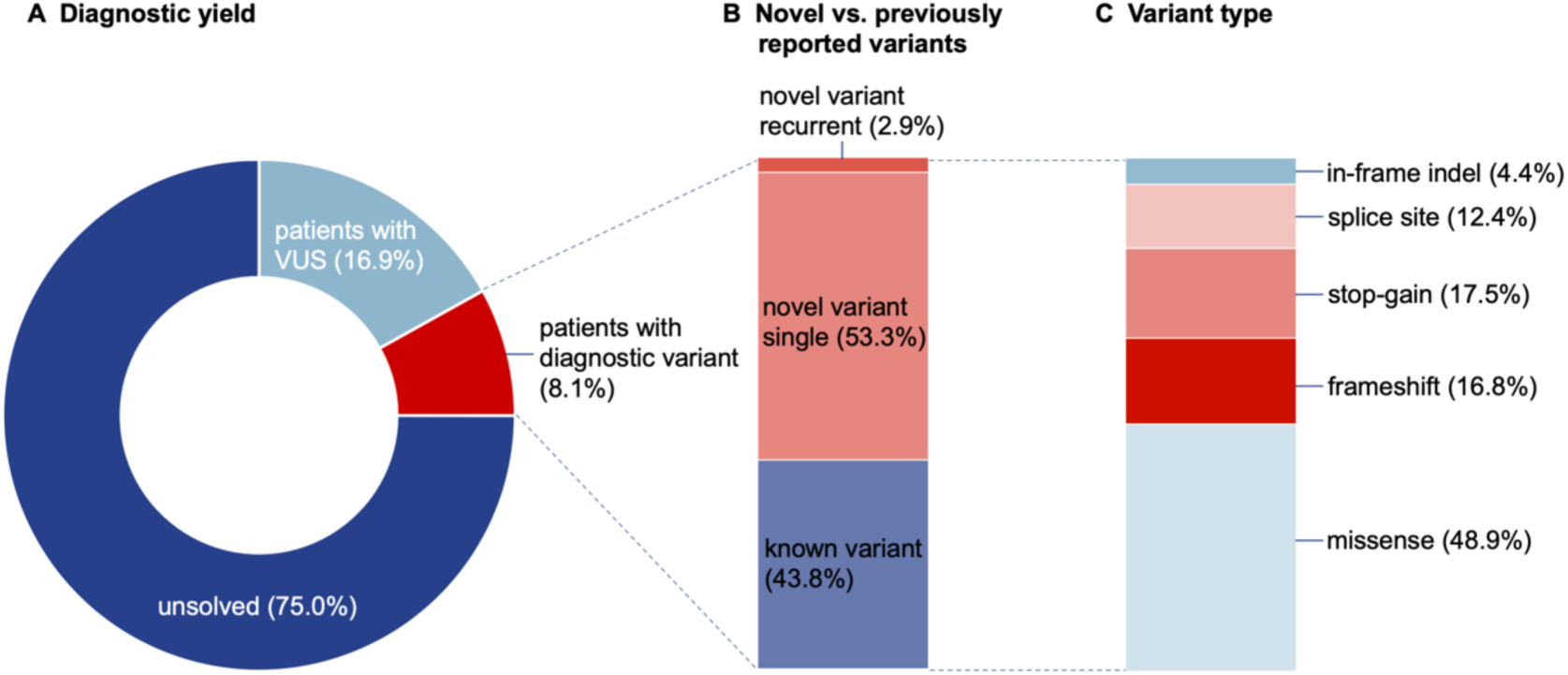
Overview of exome sequencing results. **(A)** Proportion of index patients for whom a diagnostic variant was identified, who carry a variant of uncertain significance (VUS), and who remain unsolved after searching for pathogenic variants in dystonia-linked genes. **(B)** Proportion of diagnostic variants that were previously reported (known variant), not previously reported and detected in a single patient or pedigree (novel variant single), and not previously reported and found recurrently in at least two unrelated dystonia patients (novel variant recurrent). **(C)** Distribution of diagnostic variants by variant type.

Compared to the unsolved patients and patients with VUS combined, patients with a genetic diagnosis had a significantly earlier AAO (Mann-Whitney U Test, *p*<0.0001), higher prevalence of generalized dystonia (Fisher’s exact test, *p*<0.0001), positive family history (Fisher’s exact test, *p*=0.043), and additional clinical features (Fisher’s exact test, *p*<0.0001).

Notably, there were no significant differences between the patients with VUS and the unsolved patients regarding these characteristics.

The diagnostic yield varied substantially across index patient groups, with the highest yield in patients with additional features (36.3%), followed by those with generalized dystonia (29.3%) and those with an AAO < 30 years (20.4%) (Figure 4a). ROC curves were generated to evaluate the predictive performance of four variables—AAO, positive family history, generalized dystonia, and additional features—along with a combined model incorporating all four variables (Figure 4b). The area under the curve (AUC), which measures predictive accuracy, was 0.68 for AAO, 0.67 for generalized dystonia, 0.62 for additional features, and 0.53 for family history. The combined model, incorporating all four predictors, achieved an AUC of 0.76, indicating the best overall predictive performance, with AAO and generalized dystonia being the strongest individual predictors.

### Genetic findings

Among the dystonia index patients with a genetic diagnosis (*n*=153, 8.1%), 137 distinct variants were identified in 51 genes previously linked to dystonic syndromes (Supplementary Table 2, Figure 2). These variants included 67 missense changes, 24 stop-gain variants, 23 frameshift variants, 17 splice site variants, and six in-frame deletions or insertions (Figure 1C and 2). The majority (132/137, 96.4%) of variants were found in the heterozygous state in genes known to be linked to dominant inheritance. One variant (in *MECP2*) was found in the hemizygous state in a male patient, associated with X-linked inheritance, and four variants (two in *GCH1* and one each in *SETX* and *SPR*) were biallelic and linked to autosomal recessive disorders. At least six variants occurred *de novo* (in *ADCY5*, *GNB1*, *IR2BPL, KCNN2*, *KMT2B*, and *VPS16*) (Supplementary Figure 1). Over half of the identified presumably pathogenic variants (77/137; 56.2%) were not previously reported in ClinVar or MDSGene.^21^. Among these, four recurred in at least two unrelated index patients (in *EIF2AK2*, *KCNMA1*, *SLC2A1*, and *VPS16*; see below), supporting pathogenicity. Details about the variants, pathogenicity scoring, and clinical information can be found in Supplementary Table 2.

**Figure 2.**
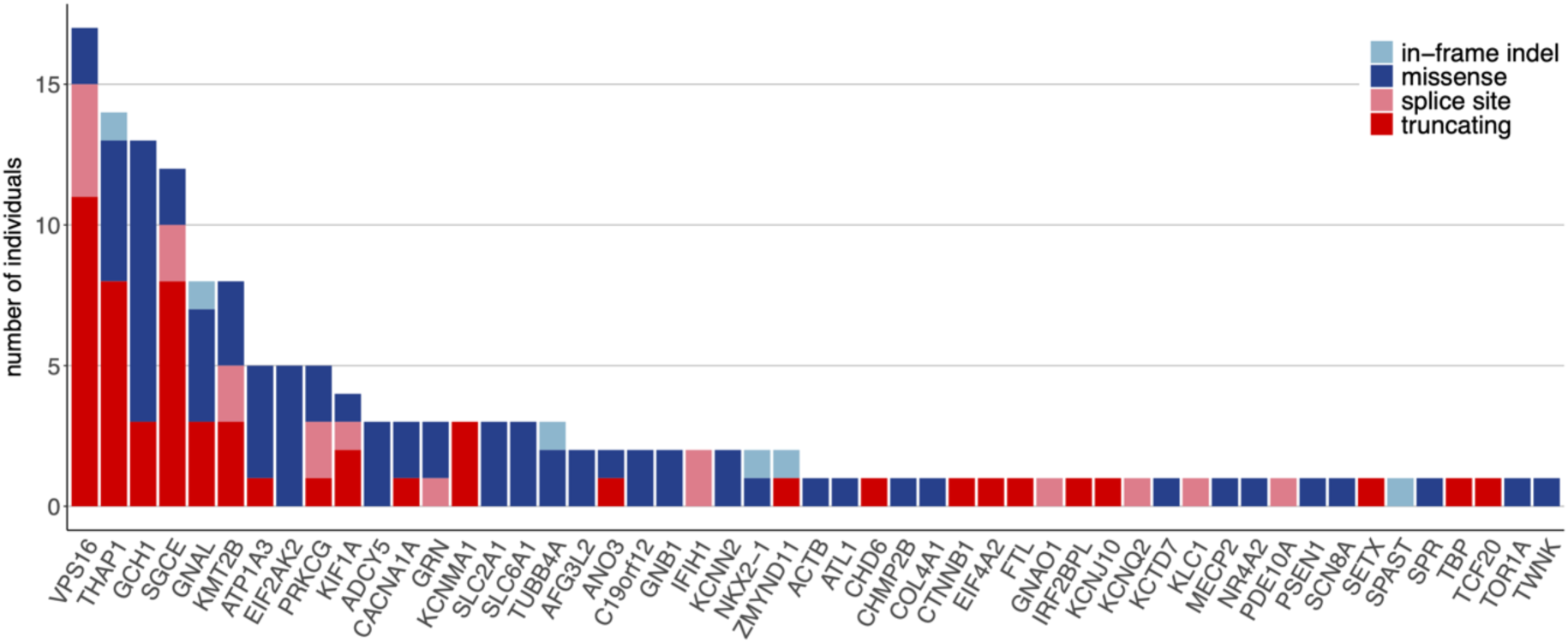
Genetic landscape in our dystonia sample (*n*=1,895 index patients) identified by exome sequencing. The number of individuals harboring a presumably pathogenic variant in genes previously linked to dystonia is shown, representing 51 distinct genetic forms and a total diagnostic yield of 8.1%. Truncating variants include stop-gain and frameshift variants. All variants were found in the heterozygous state and affect genes known to be associated with dominant inheritance, except for two homozygous variants in *GCH1*, one homozygous variant in *SPR* and *SETX*, respectively, and one hemizygous variant in *MECP2*.

A wide spectrum of genetic causes was identified, encompassing established dystonia genes as defined by the MDS Nomenclature Task Force in 103 out of 163 diagnoses, movement disorder or developmental delay genes that frequently include dystonia as a phenotype (9/163 and 4/163 diagnoses, respectively), genes typically linked to other neurological disorders in which dystonia has been rarely reported (41/163 diagnoses), and dystonia candidate genes (7/163 diagnoses) (Supplementary Table 2). Genotype-phenotype relationships were consistent with the literature in most patients (111/163; 68.1%), partially consistent in 14 (8.6%), quite inconsistent in 25 (15.3%), and unclear/unknown in 14 (8.6%) (Supplementary Table 2). Notably, all variants in established dystonia genes demonstrated at least partial genotype-phenotype consistency. Meanwhile, cases classified as "quite inconsistent" were mostly focal (*n*=15) or segmental/multifocal (*n*=8) dystonia and involved variants in genes typically associated with other disorders where dystonia is rarely reported, such as *C19orf12*, *GRN*, *KIF1A*, and *PRKCG*.

#### Findings in established dystonia genes

The most frequently implicated genes included the well-established isolated and combined dystonia genes *VPS16* (*n*=17 index patients), *THAP1* (*n*=14), *GCH1* (*n*=13), *SGCE* (*n*=12), *GNAL* (*n*=8), and *KMT2B* (*n*=8). Other dystonia genes with likely pathogenic or pathogenic variants were *EIF2AK2* (*n*=5 index patients), *ANO3* (*n*=2), *EIF4A2* (*n*=1) and *TOR1A* (*n*=1) for genes causing isolated dystonia. Of these, the genes *VPS16*, *EIF2AK2*, and *EIF4A2*, which were relatively recently associated with isolated dystonia, were not included in the prescreening process. The frequencies of these genetic forms in our dystonia sample were 0.9% (17/1,895 index patients) for *VPS16*, 0.3% (5/1,895) for *EIF2AK2* and 0.05% (1/1,895) for *EIF4A2*. Additionally, likely pathogenic or pathogenic variants were found in *ATP1A3* (*n*=5) and *GNAO1* (*n*=1) for genes causing combined dystonia, as well as *ACTB* (*n*=1), *IRF2BPL* (*n*=1), *SPR* (*n*=1), and *TUBB4A* (*n*=3) for genes linked to dystonia with other neurological or systemic features (Supplementary Table 2). Notably, 43 out of 84 variants (51.2%) in established dystonia genes were not previously reported, including two novel recurrent variants (*EIF2AK2*:p.Pro31Ser and *VPS16*:p.Ile484Thrfs*70).

For six variants, segregation analysis revealed that the identified variant co-segregated with the dystonia phenotype in the affected families, supporting its pathogenic role (*EIF2AK2*:p.Pro31Ser, *EIF2AK2*:p.Gly130Arg, *THAP1*:p.Ser51Arg, *GCH1*:p.Glu61*, *SGCE*:p.Met174Lys, *SGCE*:c.233-1G>A:p.?) (Supplementary Figure 1). Additionally, three variants were confirmed to be *de novo* (*KMT2B*:p.Arg1771Trp, *VPS16*:p.Lys397Glu, and *IRF2BPL*:p.Gln167*).

Of note, for several established dystonia genes, no carrier of a disease-causing variant was identified in our large patient group (e.g., in *AOPEP*, *HPCA*, *PRKRA*, and *TH*).

Interestingly, one patient with early-onset generalized dystonia harbored two known, at least likely pathogenic variants in different genes (*TOR1A*:p.Arg288Gln and *SPAST*:p.Glu418del).

For 32 rare variants in *KMT2B*, identified in 45 patients, the disease-specific methylation pattern (“episignature”) in patients’ blood was analyzed to assess their functional effect. Two of the tested variants were shown to result in strong hypermethylation and showed mean(z) and CV values characteristic of loss of KMT2B function (Supplementary Table 4), which was interpreted as positive functional evidence during pathogenicity scoring. Notably, these two variants were canonical splice site variants (c.4779+1G>A and c.3789+1G>A), not previously reported, and associated with childhood-onset generalized dystonia (Supplementary Table 2). Family history was negative in both patients; however, family members were not available to test whether the variants arose *de novo*.

### Findings in movement disorder genes that frequently include dystonia

Findings in movement disorder genes that frequently include dystonia as a phenotype included three patients with variants in *ADCY5*, three in *SLC2A1*, two in *C19orf12*, and one in *FTL*. This encompassed recurrent variants in *ADCY5*, *C19orf12*, and *SLC2A1*. Two unrelated patients with a mixed movement disorder phenotype, including early-onset generalized dystonia, carried the same known pathogenic variant in *ADCY5* (p.Arg418Trp), which was proven to be *de novo* in one case. A previously reported missense variant in *C19orf12* (p.Gly58Arg) was identified in two unrelated patients with adult-onset focal dystonia: one presented with cervical dystonia and the other with blepharospasm, both without additional features. For *SLC2A1*, two unrelated patients carried the same novel missense change (p.Gln25Lys), both suffering from adult-onset cervical dystonia, with one also having upper limb and shoulder dystonia. Furthermore, a patient with adult-onset isolated cervical dystonia carried a known pathogenic variant in the *FTL* gene (p.Glu58*).

### Findings in neurodevelopmental delay genes that frequently include dystonia

Presumably pathogenic variants in neurodevelopmental disorder genes that frequently include dystonia as a phenotype were identified. These included a novel nonsense variant in *CTNNB1* (p.Gln4*) in a patient with cervical dystonia without any additional features, a recurrent *de novo* missense variant in *GNB1* (p.Lys337Gln) in two unrelated patients with infancy-onset generalized dystonia - one of whom had a complex phenotype including global developmental delay, hypotonia, myoclonus, and vertical supranuclear gaze palsy, and was recently published^22^ - and a known hemizygous missense variant in *MECP2* (p.Arg97Cys) in a male patient with cervical and truncal dystonia, along with developmental delay (Supplementary Table 2).

#### Findings in genes usually linked to non-dystonia phenotypes

We identified 41 dystonia patients with likely pathogenic or pathogenic variants in 23 uncommonly dystonia-linked genes. Among these, seven showed a consistent genotype-phenotype relationship, four had partial consistency, 21 were quite inconsistent, and nine were unclear/unknown (Supplementary Table 2). While 13 of the variants had been reported in the context of other phenotypes, 24 of these variants were previously undescribed.

Genes with pathogenic variants in multiple affected patients included genes associated with spinocerebellar ataxia (*PRKCG* (*n*=5), *CACNA1A* (*n*=3), and *AFG3L2* (*n*=2)), spastic paraplegia (*KIF1A* (*n*=4)), frontotemporal dementia (*GRN* (*n*=3)), paroxysmal movement disorders (*KCNMA1* (*n*=3)), and others *(SLC6A1* (*n*=3), *IFIH1* (*n*=2), and *NKX2-1* (*n*=2)), while only one carrier of a presumably pathogenic variant was found in 14/23 uncommon dystonia genes (in *ATL1*, *CHMP2B*, *COL4A1*, *KCNJ10*, *KCNQ2*, *KCTD7*, *PDE10*, *PSEN1*, *SCN8A*, *SETX*, *SPAST*, *TBP*, *TCF20*, and *TWNK*).

Notably, four variants were recurrently found in at least two unrelated patients (*AFG3L2*:p.Arg280Trp, *GRN*:p.Cys139Arg, *IFIH1*:c.454-1G>T, *KCNMA1*:p.Ser11Alafs*15).

Except for one patient with the *AFG3L2* variant, these showed quite inconsistent or unclear genotype-phenotype relationships, i.e., in our study, were identified in patients with isolated, persistent dystonia, but are otherwise usually reported in conjunction with other phenotypes.

#### Findings in dystonia candidate genes

We also investigated dystonia candidate genes, uncovering seven novel likely pathogenic variants that were all absent from control databases and predicted to be deleterious (Supplementary Table 2). This included two missense variants in *KCNN2*, with one (p.Leu611Ile) occurring *de novo* in a patient with infancy-onset generalized myoclonus-dystonia who also showed signs of bradykinesia and ataxia.

Two variants were identified in *ZMYND11*, including a stop-gain variant (p.Arg495*) in a patient with infancy-onset generalized dystonia and an in-frame insertion (p.His315dup) in a patient with cervical dystonia, tremor, and polyneuropathy with unknown AAO. One variant each was identified in the genes *CHD6* (p.Glu2697Thrfs*29 in a patient with adolescence-onset generalized dystonia with myoclonus and ataxia), *KLC1* (c.*1+1G>A in a patient with adult-onset cervical dystonia), and *NR4A2* (p.Gln273Arg in a patient with adult-onset cervical dystonia). For *KLC1* and *NR4A2*, four additional patients each with similar phenotypes of adult-onset focal dystonia carried VUS (Supplementary Table 3, Figure 3).

**Figure 3.**
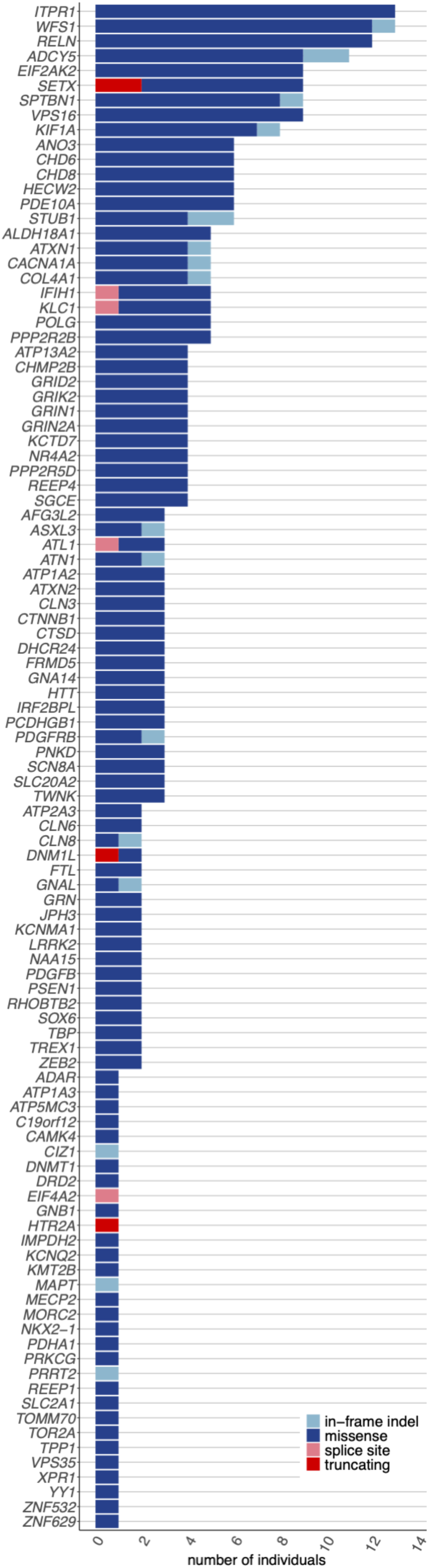
Variants of uncertain significance in our dystonia sample (*n*=1,895 index patients) identified by exome sequencing. The number of individuals harboring variants of uncertain significance in genes previously linked to dystonia is shown. This represents 329 distinct variants in 102 genes found in 321 index patients (16.9%). Truncating variants include stop-gain and frameshift variants. All variants were found in the heterozygous state and affect genes associated with dominant inheritance, except for one hemizygous variant in *MECP2* and *PDHA1,* respectively.

**Figure 4.**
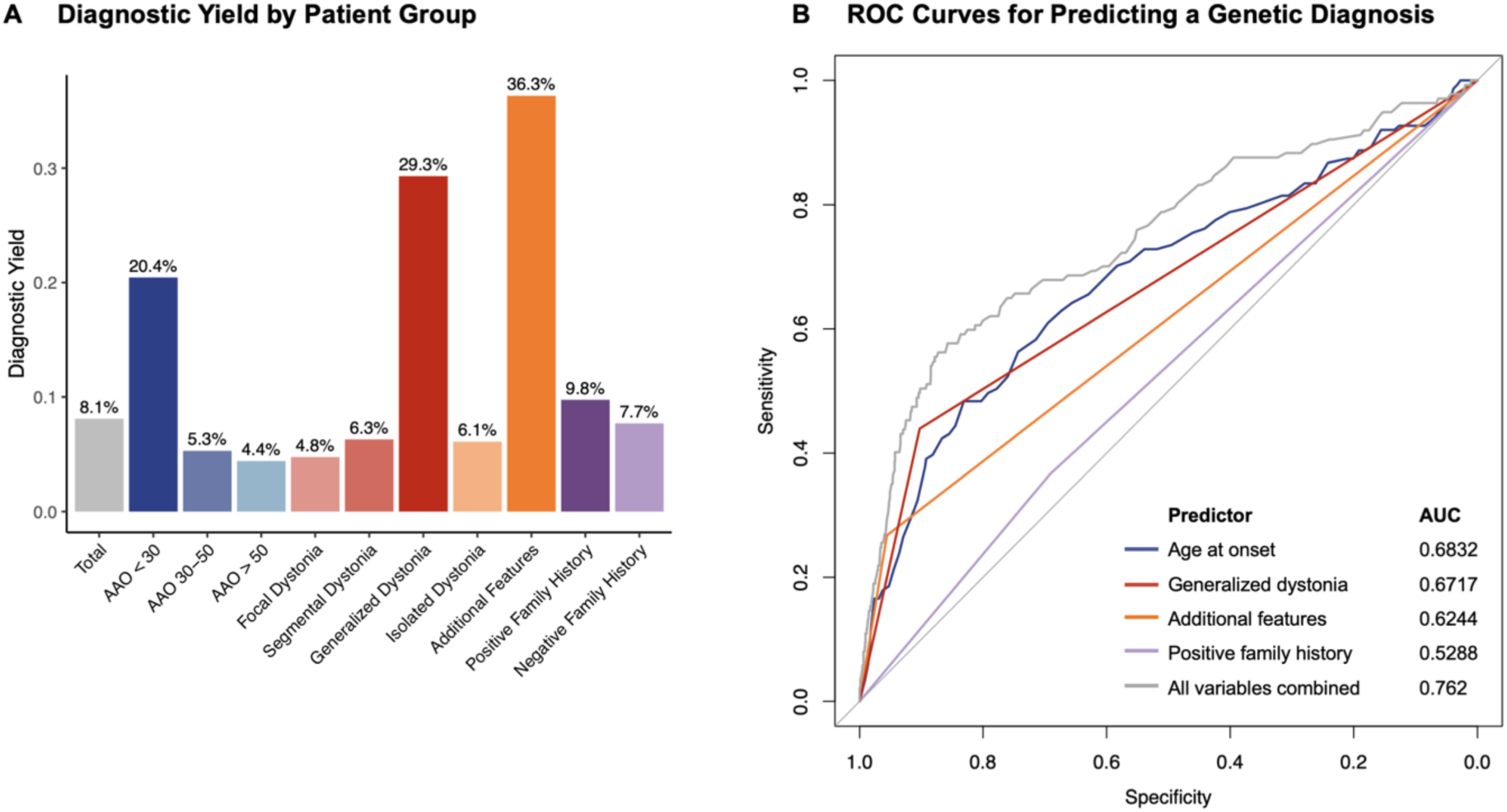
Diagnostic yield across patient groups and predictive performance of clinical factors for genetic diagnosis. **(A)** Bar plot showing the diagnostic yield (%) across different patient groups, considering index patients only. The segmental dystonia group includes patients with multifocal dystonia. **(B)** ROC curves illustrating the performance of individual predictors and a combined model for genetic diagnosis. The predictors include age at onset, generalized dystonia, additional features, positive family history, and a combined model incorporating all four predictors. Each curve shows the balance between sensitivity (true positive rate) and 1-specificity (false positive rate), with the Area Under the Curve (AUC) indicating predictive performance.

No carriers of presumably pathogenic variants in other dystonia candidate genes were detected among our 1,924 patients, including the recently proposed genes *ATP5F1B*, *ACBD6*, and *SPTBN1*. However, for *SPTBN1*, nine patients were found to carry rare missense and in-frame indels classified as VUS. They all shared an adult-onset phenotype of focal or segmental dystonia involving the upper body (Supplementary Table 3).

## Discussion

This study comprehensively examined the genetic landscape of dystonia by evaluating a cohort of 1,924 dystonia patients, most of whom (94%) had isolated dystonia. To our knowledge, to date, this is the largest next-generation sequencing study conducted in dystonia patients, providing valuable insights into the frequency and genetic diversity of dystonia-linked genes and their associated phenotypes. Despite largely excluding patients with positive prescreening for known pathogenic variants, the diagnostic yield of the present study was 8.1% (153/1,895 index patients), with an additional 16.9% of patients harboring VUS. This dataset offers crucial information on the relevance of different genetic forms of dystonia and will aid future variant interpretation and clinical diagnostics. It also highlights the weaknesses of diagnostic panels, as pathogenic variants may still escape their identification, and future re-analyses for new dystonia genes are not possible, in contrast to exome sequencing. Presumably pathogenic variants include 77 (56.2%) novel variants, with recurrent variants in *EIF2AK2*, *VPS16*, *KCNMA1*, and *SLC2A1*, and novel variant types such as two splice site variants in *KMT2B*, supported by functional evidence. For *KMT2B*, we conducted episignature analysis, which has proven to reliably distinguish between benign and pathogenic variants.^16^ An overview of the results of episignature testing has also been added to the MDSGene website (https://www.mdsgene.org), where it currently includes data on 100 distinct variants in *KMT2B*.

Patients with diagnostic variants had a significantly lower median AAO and a higher prevalence of generalized dystonia, additional neurological or systemic features, and positive family history compared to those without a diagnosis. ROC curve analysis indicated that AAO and the presence of generalized dystonia were the strongest predictors of a genetic diagnosis and diagnostic yield were substantially higher in these two subgroups (yield in generalized dystonia: 28.6%, AAO<30: 20.4%). These findings align with previous research and highlight the importance of considering exome sequencing for patients with early disease onset and generalization.^4,23^ The diagnostic yield in genetic studies of dystonia is markedly influenced by patient selection criteria, as demonstrated by reported yields ranging from 11.7% to 37.5% in different studies.^24^ For instance, a previous exome sequencing study, involving 764 dystonia patients, reported a diagnostic yield of 19%; however, the diagnostic yield reached almost 50% in patients with early-onset (<20 years), generalized, non-isolated dystonia, while it was as low as 1% in patients with late-onset, isolated, focal dystonia.^4^

### Genotype-phenotype relationships and uncommon findings

The genetic causes identified in this study are diverse, as evidenced by the detection of presumably pathogenic variants in 51 genes, highlighting the significant genetic heterogeneity observed in dystonia. Findings encompassed variants in established dystonia genes as defined by the MDS Nomenclature Task Force (62.6% of diagnoses), movement disorder genes or neurodevelopmental delay genes that frequently include dystonia as a phenotype (5.5% and 2.5%), genes typically linked to other neurological disorders where dystonia has been rarely reported (25.2%), and dystonia candidate genes (4.3%).

For the majority of variants found in isolated dystonia genes, the patients’ phenotypes generally aligned with the phenotypic spectrum described in the literature for the respective genes, thus, the genotype-phenotype relationships were classified as consistent (Supplementary Table 2). However, some unusual findings were also observed.

One patient with DYT-EIF2AK2 featured bradykinesia and rigidity, symptoms not previously reported in this form.^12,21^ A patient with DYT-ANO3 presented with spasticity and parkinsonism, features described only twice in the literature,^11,21^ suggesting they may be recurrent characteristics. Bradykinesia and resting tremor were observed in a DYT-VPS16 patient; while resting tremor has been previously reported in one patient,^25^ presence of bradykinesia is novel. Additionally, neurodevelopmental delay was noted in another DYT- VPS16 patient, which had not been previously described. Furthermore, chorea was observed in a DYT-VPS16 patient, a feature recently reported in a single patient,^26^ highlighting that additional features are not uncommon in this genetic form despite isolated dystonia cases predominating. We also identified two DYT-VPS16 patients with unusually late ages at onset (in their 60s or 70s). The reported onset of DYT-VPS16 ranges from 3 to 50 years.^21^ The occurrence of both very early-onset and these exceptionally late-onset cases, both caused by truncating mutations, suggests that additional genetic or environmental factors contribute to the variability in severity observed in DYT-VPS16. Of note, reduced penetrance, which can be regarded as extremely late age at onset, has also been reported for DYT-VPS16.^27^

Findings in combined dystonia genes encompassed *ATP1A3*, *GCH1*, *GNAO1*, and *SGCE*, generally aligning with the phenotypic spectrum described in the literature for each gene. Unusual presentations included a DYT/PARK-GCH1 patient with macrocephaly in addition to dopa-responsive dystonia. However, the possibility that the unusual additional phenotype is caused by another genetic variant (dual molecular diagnosis) cannot currently be excluded. Among the 18 DYT/MYC-SGCE patients, 13 exhibited typical myoclonus-dystonia phenotypes, whereas in the remaining five, myoclonus was either absent or not clearly documented. *SGCE* variants can also manifest as isolated dystonia, albeit rarely.^21^ *GNAO1* variants are typically linked to severe combined chorea-dystonia, but recent studies have identified splice site variants and haploinsufficiency linked to milder phenotypes and isolated dystonia.^28,29^ Interestingly, in our study, a patient with a novel splice site variant presented solely with adolescence-onset cervical dystonia, consistent with these recent observations.

Variants in *ATP1A3* show a broad phenotypic spectrum, complicating genotype-phenotype evaluations. Two of our five patients experienced a rapid onset of dystonic symptoms in adolescence, a known presentation for *ATP1A3*, although typically accompanied by parkinsonism.^13^ One of these patients also had deafness, often associated with *ATP1A3* in the context of CAPOS syndrome (Cerebellar ataxia, areflexia, pes cavus, optic atrophy, and sensorineural hearing loss).^30^ Notably, this patient carried the same variant (p.Glu818Lys) previously described in CAPOS. Four of the five patients had isolated dystonia. This includes a patient with hemidystonia and the known pathogenic variant p.Thr613Met, which has only been associated with rapid-onset dystonia-parkinsonism until now.^31^ These findings suggest that isolated dystonia can be part of the broad phenotypic spectrum of *ATP1A3*.

Findings in genes linked to dystonia with additional neurological or systemic features included *ACTB*, *IRF2BPL*, *SPR*, and *TUBB4A*, with phenotypes generally aligning with previous reports. One case involved generalized dystonia and deafness due to a known *ACTB* variant (p.Arg183Trp), while another patient with a *de novo* frameshift variant in *IRF2BPL* (p.Gln167*) exhibited generalized dystonia, learning disability, spasticity, and gait dysfunction. Additionally, a known homozygous missense variant in *SPR* (p.Arg150Gly) was linked to infancy-onset, dopa-responsive upper limb dystonia, along with depression and an anxiety-related disorder. Although *SPR* is typically associated with dystonia, parkinsonism, and developmental delay, isolated dystonia has been observed in 3% of cases,^32^ predominantly affecting the upper limbs, consistent with our patient’s presentation. *TUBB4A* variants are associated with a broad phenotypic spectrum, including isolated dystonia, typically laryngeal, cervical, or upper limb dystonia, ^33^ as seen in three patients in this study.

As new discoveries unfold, understanding the correlations between clinical manifestations and genetic variants has become increasingly complex. Mechanisms such as variable expressivity, incomplete penetrance, and genetic pleiotropy contribute to this complexity. As anticipated, our study revealed instances where the relationship between genotype and phenotype was quite inconsistent (25 out of 163 diagnoses, 15.3%). This included heterozygous variants in the genes *AFG3L2*, *ATL1*, *C19orf12*, *CHMP2B*, *COL4A1*, *GRN*, *KCNMA1*, *KCTD7*, *KIF1A*, *PDE10A*, *PRKCG*, *PSEN1*, *SCN8A*, *SLC2A1*, and *SLC6A1*. For some of these genes, further investigations are warranted to evaluate their true pathogenicity, as they are not backed by enough evidence to postulate a phenotypic expansion. However, some findings are particularly interesting:

Three patients carried variants in the frontotemporal dementia gene *GRN* (a recurrent missense and a splice site variant) that have all been previously described in frontotemporal dementia patients. In contrast to previous reports, all our patients had adult-onset focal dystonia (cervical or upper limb) without additional features to date (AAE: 25-76 years). Remarkably, previous reports describe phenotypic differences even within the same family,^35^ and the splice site variant found in our patient was previously reported in a patient with corticobasal syndrome in combination with focal dystonia.^34^ This suggests that dystonia may occur without or prior to cognitive and behavioral symptoms due to *GRN* variants.

We found three patients with novel frameshift variants in *KCNMA1*, including a recurrent variant in two unrelated individuals. *KCNMA1* is typically associated with paroxysmal non-kinesigenic dyskinesia, developmental delay, and seizures, with LoF being a known disease mechanism.^36^ The three patients presented with childhood- or adult-onset focal persistent dystonia (cervical or upper limb) without additional features, except for tremor and left eye ophthalmoplegia in one patient. Although movement disorders are a consistent feature of *KCNMA1*-related disease, particularly in LoF variants,^36^ this is the first report of isolated dystonia. Three high-confident LoF variants, including a recurrent one, were identified, suggesting that *KCNMA1* may present with isolated dystonia.

We identified four likely pathogenic variants in *KIF1A*, including two truncating variants, one splice site variant, and one missense variant affecting the kinesin motor domain, where the majority of pathogenic variants are found.^37^ Although *KIF1A* is typically associated with spastic paraplegia, the patients in this study presented with isolated upper body dystonia (cranial, cervical, or upper limbs), with two patients exhibiting tremor and one showing proximal muscle atrophy at the shoulders. While dystonia is not uncommon in *KIF1A*-related diseases, this is the first report of patients without spasticity, potentially expanding the known phenotypic spectrum.

Another interesting gene was *CACNA1A*, which has been linked to several diseases, including episodic ataxia, cerebellar ataxia, and developmental and epileptic encephalopathy. Recent reports show that dystonia can also be primary and/or generalized in cases with *CACNA1A*- variants.^38–40^ In our study, we identified three patients with likely pathogenic variants in *CACNA1A*, all of whom had isolated focal or segmental dystonia affecting the cranial and/or cervical muscles, with disease onset in their 40s. This confirms that variants in *CACNA1A* are also linked to isolated dystonia.

### Dystonia candidate genes

We also screened our data for proposed dystonia candidate genes, identifying presumably pathogenic variants in *CHD6*, *KCNN2*, *KLC1*, *NR4A2*, and *ZMYND11*, but not in others, e.g., *ATP5F1B*, *ACBD6*, *CIZ1*, *SHQ1* and *SPTBN1*.

Variants in *KCNN2* were initially associated with neurodevelopmental disorders, sometimes accompanied by movement disorders.^41^ Meanwhile, they were linked to tremulous myoclonus-dystonia,^42^ currently awaiting replication as a myoclonus-dystonia gene.^7^ We here identified two patients with likely pathogenic missense variants. The first patient had infancy-onset generalized dystonia, myoclonus, ataxia, and bradykinesia – symptoms previously reported in *KCNN2*-related disease^41^ - and carried a *de novo* variant, supporting its pathogenicity and confirming the role of *KCNN2* in myoclonus-dystonia.

The second missense variant (p.Ser376Leu) is located in the ion channel domain of the protein, which is highly intolerant to missense variants^43^ and harbors other pathogenic variants.^41,42^ This patient has had cervical dystonia since the late 20s, without additional features reported, which only partially aligns with previous findings and may broaden the phenotypic spectrum of *KCNN2* to include later-onset isolated dystonia.

In 2020, two missense variants in *CHD6* were linked to infancy-onset generalized dystonia and developmental delay upon exome sequencing.^4^ We also identified a novel frameshift variant in a patient with adolescence-onset generalized dystonia with myoclonus and ataxia. This variant was classified as likely pathogenic due to its absence in control databases and *CHD6*’s intolerance to LoF variants (pLI-score of 1). The same study also identified a presumably disease-causing missense variant in *KLC1* in a patient with childhood-onset generalized dystonia and developmental delay.^4^ In our sample, we found a novel splice site variant in *KLC1* in a patient with late-onset cervical dystonia with no reported additional features, as well as five missense VUS in patients with adult-onset upper limb dystonia. Additionally, *ZMYND11* was proposed as a cause of childhood-onset focal dystonia and intellectual disability.^4^ In our study, we identified two patients with presumably pathogenic variants in *ZMYND11*: a single amino acid duplication in a patient with cervical dystonia, tremor, and polyneuropathy of unknown AAO, and a stop-gain variant in a patient with infancy-onset generalized dystonia with no additional features reported.

Although no pathogenic variants were identified in the recently proposed candidate gene *SPTBN1*, we discovered nine patients with rare missense or in-frame VUS. The affected patients shared adolescence or adult onset (median AAO: 40 years, range: 16-46) focal or segmental dystonia that mainly affected the neck or upper extremities. In contrast, the initial report described a splice site variant in a patient with adolescent-onset segmental dystonia and developmental delay.^4^

Furthermore, we identified a novel, likely pathogenic missense variant in *NR4A2*, a gene recently associated with levodopa-responsive dystonia and developmental delay.^44,45^ Notably, our patient’s presentation was primarily characterized by cervical dystonia with no additional features reported, and information on levodopa responsiveness was unavailable. Additionally, four other patients harbored missense variants in *NR4A2* classified as VUS, all located within the ligand-binding domain at locations intolerant to missense changes.^43^ These patients all presented with adult-onset focal dystonia affecting the neck or upper limbs, with AAO ranging from 27 to 44 years.

It remains to be seen whether variants in *CHD6*, *KLC1*, *NR4A2*, *SPTBN1,* and *ZMYND11* significantly contribute to dystonia pathogenesis and if different types of variants (e.g., missense vs. truncating) are linked to varying clinical manifestations, as observed in other disorders. Our data suggest a possible role of these genes in dystonia without neurodevelopmental features.

### Limitations

Despite the strengths of our exome sequencing study in identifying pathogenic variants in a large sample of dystonia patients, there are some limitations. First, exome sequencing may miss non-coding variants and structural variations that could contribute to dystonia. Notably, whole genome sequencing is currently underway for a subset of yet-unsolved patients and may increase the diagnostic yield by 5-10%.^46^ Second, the study’s exclusion of patients with known pathogenic variants may have biased the sample towards rarer or novel genetic causes, limiting the generalizability of prevalence and observed genotype-phenotype relationships. This is underlined by the absence of the most frequent dystonia variant, the p.E303del in *TOR1A*, since this variant was excluded by prescreening. Third, the classification of VUS remains challenging, and co-segregation studies or functional studies are necessary to confirm their pathogenicity. Access to family members for segregation analysis is often limited and functional studies are currently available only for a small subset of tested genes, including *KMT2B*. Additionally, the inherent limitations of clinical phenotyping must be acknowledged. Variability in expert assessments can lead to misdiagnosis, and within this large cohort, some patients may have alternative explanations for their symptoms, such as functional neurological disorders or orthopedic causes. While AI-guided tools like video analysis or machine-learning diagnostics may reduce this variability in the future, they are not yet implemented in clinical practice.

### Conclusion and Outlook

Taken together, our study demonstrates the usefulness of exome sequencing to elucidate the molecular basis in a heterogeneous disease like dystonia. Given the considerable number of genes and variants linked to dystonia as well as the continuously growing candidate gene list, diagnostic gene-by-gene approaches and specific gene panels are often impractical, and comprehensive screening strategies, such as exome or genome sequencing, are the most efficient method to arrive at a diagnosis. These unbiased screening strategies can be reanalyzed at any time to evaluate, for example, novel disease genes and will increase the diagnostic yield in the long term. To aid future variant interpretation, we therefore also provide a list of all (*n*=321) index patients with VUS in dystonia-linked genes (Supplementary Table 3), which may be reclassified to likely pathogenic variants with future efforts, i.e., by identifying additional carriers. This is particularly important for dystonia candidate genes such as *CHD6*, *KLC1, NR4A2,* and *SPTBN1*, in which several VUS with similar phenotypic presentations were identified.

Finding a genetic diagnosis is invaluable as it facilitates clinical management, treatment decisions, and genetic counseling, provides prognostic information, and offers crucial insights for the development of targeted therapies.

## Data availability

Raw data were generated at the Competence Centre for Genomic Analysis in Kiel, Germany, and are available from the corresponding author on request.

## Acknowledgments

The authors acknowledge support from the Lübeck High Performance Compute Infrastructure “OmicsCluster”.

## Competing interests

K.E.Z. has received research support from Strathmann and the German Research Council. She reports speaker’s honoraria from Bayer Vital GmbH, BIAL, AbbVie, Alexion, Allergan and Merz outside the submitted work. She has served as a consultant and received fees from Merz, Ipsen, Alexion, Bial and the German Federal Institute for Drugs and Medical Devices (BfArM).

J.S.P. has research support RF1NS075321, RO1NS134586, RO1NS103957, NS107281, NS097437, U54NS116025, U19 NS110456, NS097799, R33 AT010753, RO1NS118146, NS124738, R01AG065214, NS124789, R21 NS133875, R21TR004422, R21TR005231.

Foundation support: Michael J Fox Foundation, Barnes-Jewish Hospital Foundation (Elliot Stein Family Fund and Parkinson disease research fund), American Parkinson Disease Association (APDA) Advanced Research Center at Washington University, Missouri Chapter of the APDA, Paula and Rodger Riney Fund, Jo Oertli Fund, Huntington Disease Society of America, Murphy Fund, Fixel Foundation, N. Grant Williams Fund, Pohlman Fund, CHDI and Prilenia. He is also co-director for the Dystonia Coalition, which has received the majority of its support through the NIH (grants NS116025, NS065701 from the National Institutes of Neurological Disorders and Stroke TR 001456 from the Office of Rare Diseases Research at the National Center for Advancing Translational Sciences).

R.L.B. performs botulinum toxin injections at the University of Rochester (50% effort); serves/has served on scientific advisory board for Allergan, Ipsen, Merz and Revance; receives research support Fox Foundation; NIH (NINDS, ORDR): Dystonia Coalition Projects, Site PI; Consultant and research study rater for Abbvie/Allergan; holds stock options in VisualDx; and serves/has served as an expert witness in legal proceedings including malpractice, not involving commercial entities.

J.V. has received consultancies from Medtronic, Boston Scientific, Ceregate, and Newronika. He has served on advisory boards for Medtronic and Boston Scientific. He reports speaker’s honoraria from Medtronic, Boston Scientific, Abbott, AbbVie, Bial, and Zambon outside the submitted work.

H.A.J. has active or recent grant support (recent, active, or pending) from the US government (NIH), private philanthropic organizations (Cure Dystonia Now, Lesch-Nyhan Syndrome Children’s Research Foundation), and industry (Abbvie, Addex, Aeon, Sage, Ipsen, Jazz).

H.A.J. has also served on advisory boards or as a consultant for the NIH (CREATE Bio DSMB) and industry (Abbvie, Addex, Ipsen, Merz, and Vima). He has received stipends for administrative work from the International Parkinson’s Disease and Movement Disorders Society. H.A.J. has also served on the Scientific Advisory Boards for several private foundations (Benign Essential Blepharospasm Research Foundation, Dystonia Medical Research Foundation). He also is principle investigator for the Dystonia Coalition, which has received the majority of its support through the NIH (NS116025, NS065701 from the National Institutes of Neurological Disorders and Stroke TR001456 from the Office of Rare Diseases Research at the National Center for Advancing Translational Sciences).

C.Kl. has served as medical advisor to Centogene, Retromer Therapeutics, Takeda, and Lundbeck and has received speakers’ honoraria from Bial and Desitin.

C.Ka. reports serving on advisory boards for Biogen and Roche outside of the submitted work. The other authors report no competing interests.

