## Supplementary Methods for "Genetic Diversity and Expanded Phenotypes in Dystonia: Insights from Large-Scale Exome Sequencing"

##### Table of Contents


| | |
|---|---|
| Supplementary Methods | 1-2 |
| Supplementary References | 3 |
| Supplementary Tables 1-4 | Excel File |

### Supplementary Methods

#### Exome sequencing and data processing

Genomic DNA was extracted from peripheral blood samples using the QIAamp DNA Mini Kit (Qiagen, Hilden, Germany) according to the manufacturer's protocols. The whole exome library preparation and sequencing were conducted at the Competence Centre for Genomic Analysis in Kiel, Germany, utilizing the Illumina DNA Prep with Enrichment kit (Illumina, San Diego, USA) and the IDT xGen Exome v2 baits (Integrated DNA Technologies Coralville, Iowa, USA). Sequencing was performed on an Illumina NovaSeq 6000 instrument using the S4 Flowcell and 150 bp paired-end sequencing with an approximate mean sequencing depth of 150x. Sequenced fastq reads underwent preprocessing to ensure data quality and alignment accuracy. Reads were trimmed, removing reads with fewer than 50 bases, those with excessively high CG content, and low-quality reads with a Phred score <15. Alignment to the Hg38 reference genome was performed using BWA-mem2.[1] Post-alignment processing included Picard tools for read cleaning, sorting, and duplicate marking (CleanSam, SortSam, MarkDuplicates). Mate-pair information was verified with FixMateInformation, reads were assigned to read groups with AddOrReplaceReadGroups, and reads were reordered with ReorderSam. Base quality recalibration was conducted using GATK's Base Quality Recalibration tool, utilizing the Homo_sapiens_assembly38.dbsnp138.vcf from GATK.[2]
Variant calling was performed to identify small insertions or deletions (indels) and single nucleotide variants (SNVs). DeepVariant[3] version 1.2.0 was utilized for variant calling, and GLnexus was employed to merge Variant Calling Files (VCF) into a single cohort file. Variant Calling Files (VCFs) were annotated using VEP v.103. Additional annotation information was provided, including CADD[4] (v1.5) score and annotation from gnomAD[5] exome (r2.1.1) and genome (r.3.0). Sample files were restructured using maftools for efficient handling and analysis.

##### Relatedness analysis

To identify possible duplicates and related individuals, a two-step approach was employed utilizing GRAPE[6] and NgsRelate.[7]
First, the GRAPE toolkit was used to analyze vcf files from all individuals, identifying IBD segments with the IBIS[8] algorithm and estimating relationship degrees with ERSA.[9] Pairs of individuals estimated to be related to the second degree or closer, along with a custom list of suspected relations based on clinical data or shared rare variants, advanced to the second step.
Second, BAM files were preprocessed to generate allele frequencies and genotype likelihoods using ANGSD.[10] These genotype likelihoods were compared to determine a more accurate percentage of IBD-sharing, yielding kinship coefficients and pairwise relatedness. IBS patterns were also analyzed with KING[11] to obtain R1 and R0 ratios. Individuals were categorized into zero-degree (monozygotic twins or same individual) or first-degree (parent-offspring or full siblings) relationships, and those with implausible R1 and R0 values were filtered out.

#### Episignature analysis for *KMT2B* variants

To assess the functional effect of *KMT2B* variants, the DYT-KMT2B-specific methylation pattern ("episignature"), comprising 113 specific CpG sites, was analyzed as described.[12] For this, peripheral blood methylation analysis was performed using the Illumina MethylationEPIC BeadChip.

DNA methylation (DNAm) profiling was conducted using the "Infinium MethylationEPIC" array (Illumina, Inc.). DNA extracts were diluted to approximately 50 ng/µl concentration and then subjected to bisulfite conversion with the EZ DNA Methylation Kit (Zymo Research), following the supplier's alternative incubation conditions for the Illumina Infinium MethylationEPIC Array. The converted DNA samples were then hybridized to the EPIC array and scanned on an iScan instrument (Illumina, Inc.) as per the manufacturer's instructions (Document #1000000077299v0). The raw DNAm intensities were generated using the iScan control software (v2.3.0.0; Illumina, Inc.) and exported in .idat format for further processing and analysis.

Methylation intensities were analyzed using R version 4.2.2 and the minfi package (v1.44). We identified poor-performing probes by minfi::detP and only kept samples that have a mean p-value over all probes <0.05. Stratified quantile normalization was performed for normalization. Following best practices, the dataset was cleaned by removing several poor-performing probes: Probes with detection p-value <0.01 in over 50% of samples, probes with multiple binding sites on the array, probes that multimap on different genetic regions based on bowtie2, and unreliable probes (based on[13]). During data cleaning, 10 of the 113 *KMT2B*-associated probes were filtered out (cg20042692, cg27519958, cg09512891, cg23793686, cg07579404, cg09423283, cg08894761, cg13876206, cg02413092, cg04988061).

The methylation level at each site was assessed as an M-value, which indicates the log2 ratio of the intensities of methylated and unmethylated states. Normalized methylation levels (z-values) for each CpG site in each *KMT2B*-variant carrier were derived by taking the aberration of the M-value from the mean M-value of that site in controls and dividing it by the standard deviation (SD) at that site in controls. As a control group, we used 17 DYT-SGCE patients and repeated the calculation using 38 unaffected individuals to compare the outcomes. We also repeated the calculation using all 113 CpG sites. The mean of the normalized methylation levels (mean(z)) and the coefficient of variation (CV = SD/ |mean|) were calculated for each individual.
